# Distinct neurostructural signatures of anxiety-, fear-related and depressive disorders: a comparative voxel-based meta-analysis

**DOI:** 10.1101/2021.11.02.21265836

**Authors:** Xiqin Liu, Benjamin Klugah-Brown, Ran Zhang, Jie Zhang, Benjamin Becker

## Abstract

Internalizing disorders encompass anxiety, fear and depressive disorders. While the DSM-5 nosology conceptualizes anxiety and fear-related disorders as an entity, dimensional psychopathology models suggest that generalized anxiety disorders (GAD) and major depression originate from an overarching “anxious-misery” factor whereas fear-related disorders originate from the “fear” factor. Given that a neurobiological evaluation is lacking, we conducted a comparative neuroimaging meta-analysis of gray matter volume alterations to determine common and disorder-specific brain structural signatures in these disorders. The PubMed, Web of Knowledge, and Scopus databases were searched for case-control voxel-based morphometric studies through December, 2020 in GAD, fear-related anxiety disorders (FAD, i.e., social anxiety disorders, SAD; specific phobias, SP; panic disorders, PD; and agoraphobia, AG) and major depressive disorder (MDD). Neurostructural abnormalities were assessed within each disorder group followed by quantitative comparison and conjunction analyses using Seed-based d-Mapping (SDM-PSI). GAD (9 studies, 226 patients) showed disorder-specific decreased volumes in left insula (*z*=-2.98, *p*_*FWE-corrected*_<0.05) and lateral/medial prefrontal cortex (z=-2.10, *p*_*FWE-corrected*_<0.05,) as well as increased right putamen volume (*z*=1.86, *p*_*FWE-corrected*_<0.05) relative to FAD (10 SAD, 11 PD, 2 SP studies, 918 patients). Both GAD and MDD (46 studies, 2,575 patients) exhibited decreased prefrontal volumes compared to controls and FAD. While FAD showed less robust alterations in lingual gyrus (*p* < 0.0025, uncorrected), this group presented intact frontal integrity. No shared structural abnormalities were found. Unique clinical features characterizing anxiety-, fear-related and depressive disorders are reflected by disorder-specific neuroanatomical abnormalities. Targeting the disorder-specific neurostructural signatures could improve therapeutic efficacy.

## Introduction

Anxiety disorders (AD) constitute the most prevalent diagnostic group of mental disorder and cause considerable suffering, disability and economic cost. AD comprise a group of heterogeneous disorders that share features of excessive fear and anxiety (1). Recent overarching conceptualizations based on DSM-5 propose that AD can be placed along a fear– anxiety continuum ranging from excessive fear-based responses to imminent specific threats in fear-related anxiety disorders (FAD, including social anxiety disorders, SAD; specific phobias, SP; panic disorders, PD; and agoraphobia, AG) to a rather diffuse anxious apprehension of events in anxiety-related anxiety disorders such as generalized anxiety disorder (GAD) (2). On the other hand, subcategories of AD are often highly co-morbid with each other as well as with other emotional (internalizing) disorders (3). Particularly, GAD and major depressive disorders (MDD) exhibit symptomatic overlap (e.g., negative affect, worry) (4) and common genetic factors (5). This echoes findings on psychopathological factor model in internalizing disorders indicating that both GAD and MDD originate from the higher-order “anxious-misery” or “distress” dimension, while SAD, SP, AG and PD originate from the “fear” dimension (6–8). Despite ongoing debates about the nosology of psychiatric disorders and overarching symptom-domains (9), the neurobiological substrates underlying these conceptualizations remain unclear.

Animal models and human neuroimaging have demonstrated that anxiety and fear are regulated by distinct neurobiological circuits such that fear response is mediated by central nucleus of the amygdala (CeA), and anxiety is mediated by bed nucleus of the stria terminalis (BNST) (10, 11). While the segregation has been translated into the Acute Threat (fear) and Potential Threat (Anxiety) domains of the Research Domain Criteria (RDoC) (12), accumulating findings indicate a shared neural basis anxiety, fear and general negative affect (13, 14). Shared and distinct neurobiological systems underlying these pathological domains have been revealed by functional magnetic resonance imaging (fMRI) studies comparing GAD and FAD (15, 16), as well as GAD and MDD during emotional and cognitive processing (17–19) or by case-control studies examining brain structural alterations using structural MRI (sMRI) in single diagnostic categories (20). However, results have been dependent on the specific task-domain and population examined and conclusions have been limited by small sample sizes and analytic variability (21).

One effort to address these issues is to conduct quantitative neuroimaging meta-analyses in order to synergize findings across individual studies and diagnostic entities. The meta-analytic approach previously allowed to determine consistent structural alterations within separate diagnostic groups (22–25), and results indicate indirect support for dissociable yet overlapping structural deficits between GAD, FAD and MDD. For instance, decreased volumes in medial prefrontal cortex (mPFC) and superior temporal gyrus (STG) and insula have been separately and concurrently found in GAD (22), FAD (24) and MDD (26). Two recent meta-analyses employing a transdiagnostic approach encompassing findings from GAD and FAD studies have reported structural deficits in the left inferior frontal gyrus (IFG) in mixed AD group (27, 28), suggesting a common neuroanatomical marker for AD. However, there is no meta-analyses that directly disentangle common and separable structural alterations between GAD, FAD and MDD. Against this background, the current comparative meta-analysis aimed to quantitatively determine shared and disorder-specific structural alterations in GAD, FAD and MDD, in accordance with the DSM-5 nosology and psychopathological factor model.

To this end, we performed a pre-registered whole-brain coordinate-based meta-analysis of case-control neuroimaging studies examining brain structural alterations using voxel-based morphometric (VBM) approaches in GAD, FAD (including SAD, SP, AG and PD) or MDD. To facilitate robust inference, the recently developed Signed Differential Mapping with Permutation of Subject Images (SDM-PSI, https://www.sdmproject.com/) approach was employed which – unlike previous analytic frameworks such as activation likelihood estimation (ALE) or standard SDM – capitalizes on the effect sizes and family-wise correction (FWE) for multiple comparisons thus promoting unbiased estimation and increased reliability (29). We hypothesized GMV deficits in prefrontal regions such as IFG in GAD (22) and orbitofrontal cortex (OFC) in MDD (26), and in limbic regions such as amygdala in FAD (30). In terms of meta-analyses revealing transdiagnostic neurobiological substrates for mental disorders (31, 32), we expected to find common alterations in regions engaged in the representation of negative affect such as the insula.

## 2. Methods

### 2.1 Search and Study Selection

A comprehensive literature search was conducted in PubMed, Web of Knowledge, and Scopus databases for case-control VBM studies comparing GAD, or SAD/SP/AG/PD, or MDD patients with HC through December 20, 2020 (**Figure 1**), according to the PRISMA guidelines (33). Details and search terms are provided in **Supplementary Methods**.

**Figure 1.**
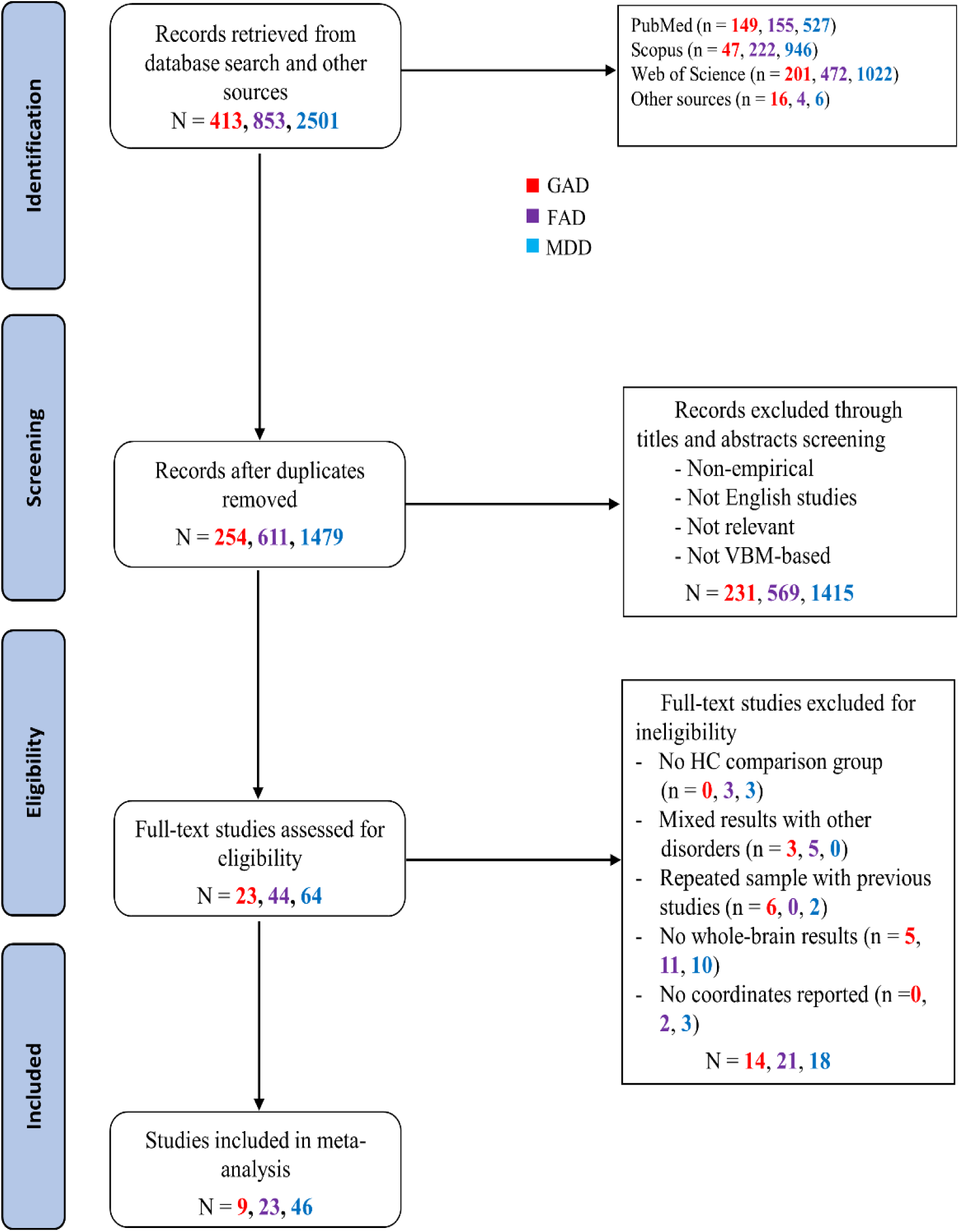
PRISMA flow diagram of study selection in the current meta-analysis. Abbreviations: FAD, fear-related anxiety disorder; GAD, generalized anxiety disorder; HC, healthy controls; MDD, major depressive disorder; VBM, voxel-based morphometry

Inclusion criteria: (1) whole-brain VBM comparisons reported between patients with a validated primary diagnosis of GAD, SAD/SP/AG/PD or MDD and HC; (2) participants < 65 years; (3) coordinates provided in Talairach or Montreal Neurological Institute (MNI) space. In case of overlapping samples between studies, only the record with the greatest sample size was included. For studies using longitudinal treatment designs, only pre-treatment (baseline) data were included. Studies of MDD patients with treatment-resistant depression were excluded because the underlying pathophysiology might be different and the brain structure might be influenced by exposure to multiple medications (34). To match illness duration between patient groups, studies of first-episodic MDD with a mean illness duration 2 years were excluded. Original whole-brain *t*-maps and missing data were requested from authors via e-mail (see **Supplementary Methods**).

X.Q.L and R.Z. independently screened and assessed all articles achieving 100% agreement. Peak coordinates and effect sizes of significant GMV alterations in both directions (i.e., patient > HC, and patient < HC) and other basic information (e.g., sample size, age, sex, etc.) were independently extracted by X.Q.L and R.Z.

### 2.2 Meta-analysis

Voxel-wise meta-analyses were performed using Seed-based d Mapping as implemented in the most recent SDM-PSI (version 6.21, https://www.sdmproject.com/), which incorporates family-wise error (FWE) correction for multiple comparisons with common permutation tests using threshold-free cluster enhancement (TFCE) (29). Original *t*-maps were included when available. Details see **Supplementary Methods**.

To determine common and disorder-specific brain structural alterations between anxiety-related, fear-related and depressive disorders, separate meta-analyses were initially conducted within each of the disorder groups (i.e., GAD vs HC, FAD vs HC, MDD vs HC) to characterize robust GMV deficits in each category. Subgroup meta-analyses in FAD were further performed to account for heterogeneity between specific diagnostic subgroups within this category. Next, disorder-specific GMV alterations were examined with quantitative contrast analyses using linear modelling and randomization tests. Lastly, shared GMV alterations between the disorders were determined with conjunction analyses for each pair of patient groups and for all three groups using a multimodal SDM tool, accounting for error in the estimation of *p*-values within each voxel from the separate meta-analytic maps.

To examine potential confounding influence of demographic and clinical variables, meta-regression analyses were performed [in case variables were reported in > 8 studies, as recommended by Radua et al. (35)] including age and female ratio in GAD, age, female ratio, illness duration, medication and comorbidity in FAD and MDD as well as symptom severity (Hamilton Depression Rating Scale, HDRS) in MDD.

A TFCE-based FWE corrected threshold *p* < 0.05 with a voxel extent ≥10 was initially used in all meta-analyses. A more liberal threshold yet balancing between Type I and Type II errors (*p* ≤ 0.0025 and voxel extent > 10 voxels) was further employed in exploratory separate meta-analyses in line with previous studies (36), and in conjunction analyses as suggested in bimodal tests (37). For the meta-regressions, a classic threshold of *p* < 0.0005 was used and only regions determined in main meta-analyses were included. Heterogeneity of significant clusters was tested with *I*^2^ statistic, with *I*^2^ < 50% indicating low heterogeneity (38). Publication bias was assessed with Egger’s test. Additionally, a meta-analysis across GAD, FAD and MDD was conducted to identify potential transdiagnostic neural markers (see **Supplementary Methods and Figure S1**).

The meta-analytic protocols were pre-registered on the Open Science Framework (https://osf.io/es2vm). Coordinates and *t*-value files are available at https://osf.io/46uc2/. Unthresholded whole-brain maps are provided at https://neurovault.org/collections/11343/.

## 3. Results

### 3.1 Included studies and sample characteristics

Included were 9 GAD studies, 23 FAD (10 SAD, 11 PD, 2 SP and 0 AG) studies, and 46 MDD studies. Whole-brain *t*-maps were available for 1 GAD study, 2 FAD (SAD) studies and 1 MDD study. **Table 1** summarizes the sample sizes (226 GAD, 918 FAD, 2,575 MDD), female ratio and age from each patient vs. HC group. Demographic and clinical information the studies in each group are presented in **Supplementary Table S1** to **S3**.

**Table 1.** Samples included in the meta-analysis.

| Study | Number of Subjects in Group |  | Female subjects, % |  | Mean Age |  | Age, SD |  |
| --- | --- | --- | --- | --- | --- | --- | --- | --- |
|  | Patients | HC | Patients | HC | Patients | HC | Patients | HC |
| **GAD** | 226 | 226 | 59 | 58 | 31.38 | 29.97 | 12.45 | 13.83 |
| **FAD** | 918 | 989 | 56 | 51 | 32.30 | 31.56 | 10.71 | 10.18 |
| SAD | 444 | 496 | 48 | 42 | 29.69 | 29.95 | 9.18 | 9.58 |
| PD | 370 | 415 | 63 | 59 | 37.01 | 34.50 | 11.17 | 10.45 |
| SP | 104 | 78 | 67 | 65 | 26.74 | 26.17 | 8.48 | 7.73 |
| **MDD** | 2575 | 2866 | 64 | 57 | 38.62 | 37.42 | 14.19 | 14.12 |

Abbreviations: GAD, generalized anxiety disorder; FAD, fear-related anxiety disorder; HC, healthy control; MDD, major depressive disorder; PD, panic disorder; SAD, social anxiety disorder; SD, standard deviation; SP, specific phobia.

Comparing age and female ratio of the three patient groups with sample size-weighted one-way ANOVA revealed significant differences in mean age (*F*_2,75_ = 3 85, *p* < 0.05; *η*^2^ = 0.09) and a marginal significant difference in female ratio (*F*_2,75_ = 2.98, *p* = 0.057, *η*^2^ = 0.07). Post-hoc tests revealed that both mean age and female ratio of MDD patients were higher than those of the FAD (*ps* < 0.05). Age and sex were consequently included as covariates in the quantitative comparative meta-analyses.

### 3.2 Regional GMV alterations

#### 3.2.1 GAD patients versus HC

Relative to HC, GAD demonstrated robust GMV decreases in the left Rolandic operculum/insula/STG and left inferior frontal gyrus (IFG). No clusters of increased GMV were found (**Table 2, Figure 2A)**. When applying a more liberal threshold (*p* < 0.0025, uncorrected), decreased GMV was found in the left insula/Rolandic operculum/STG, left IFG, left thalamus, right lingual gyrus and right inferior parietal gyrus (IPG), while increased GMV was present in the right paracentral lobule (see **Table S4, Figure 2A**).

**Table 2.**
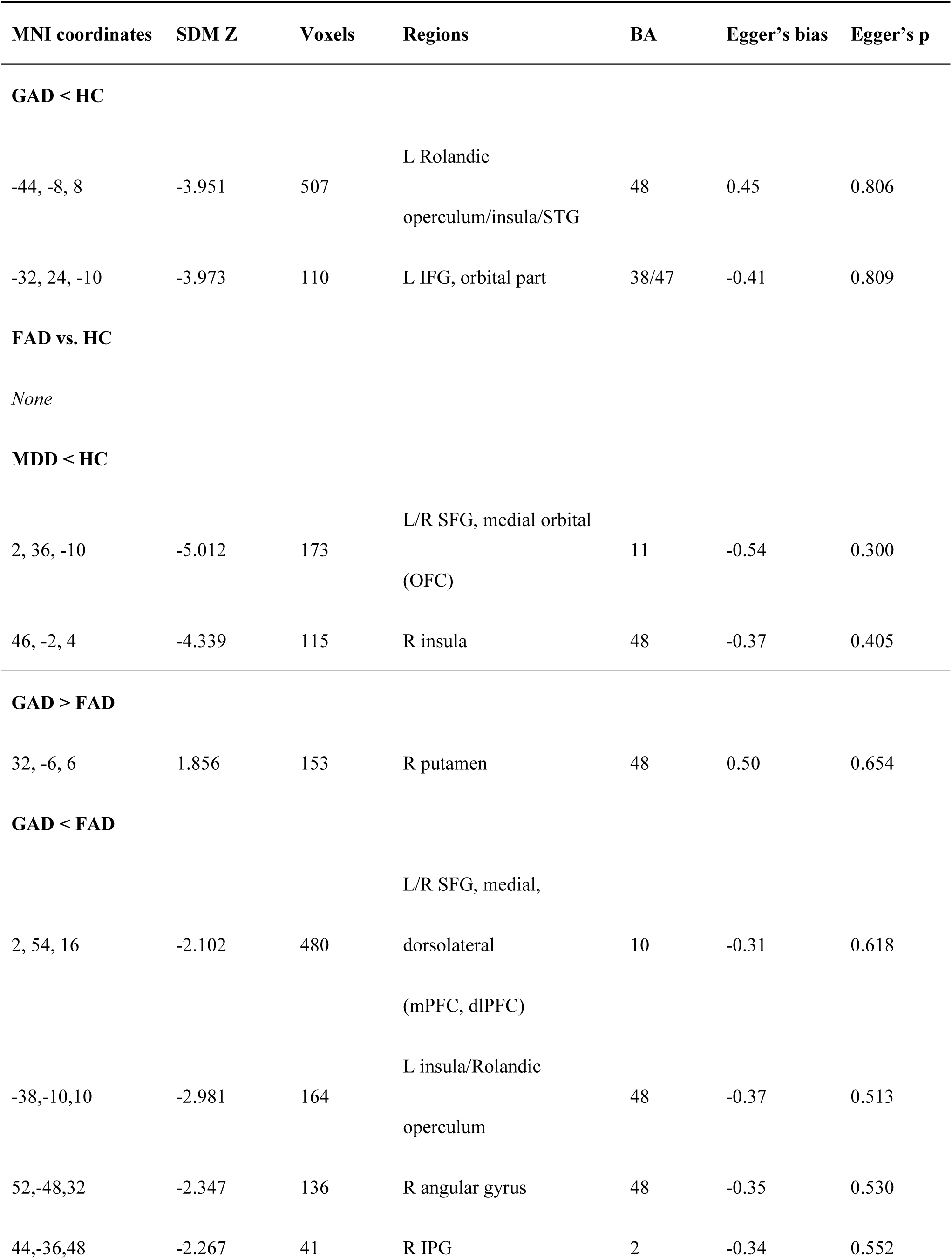
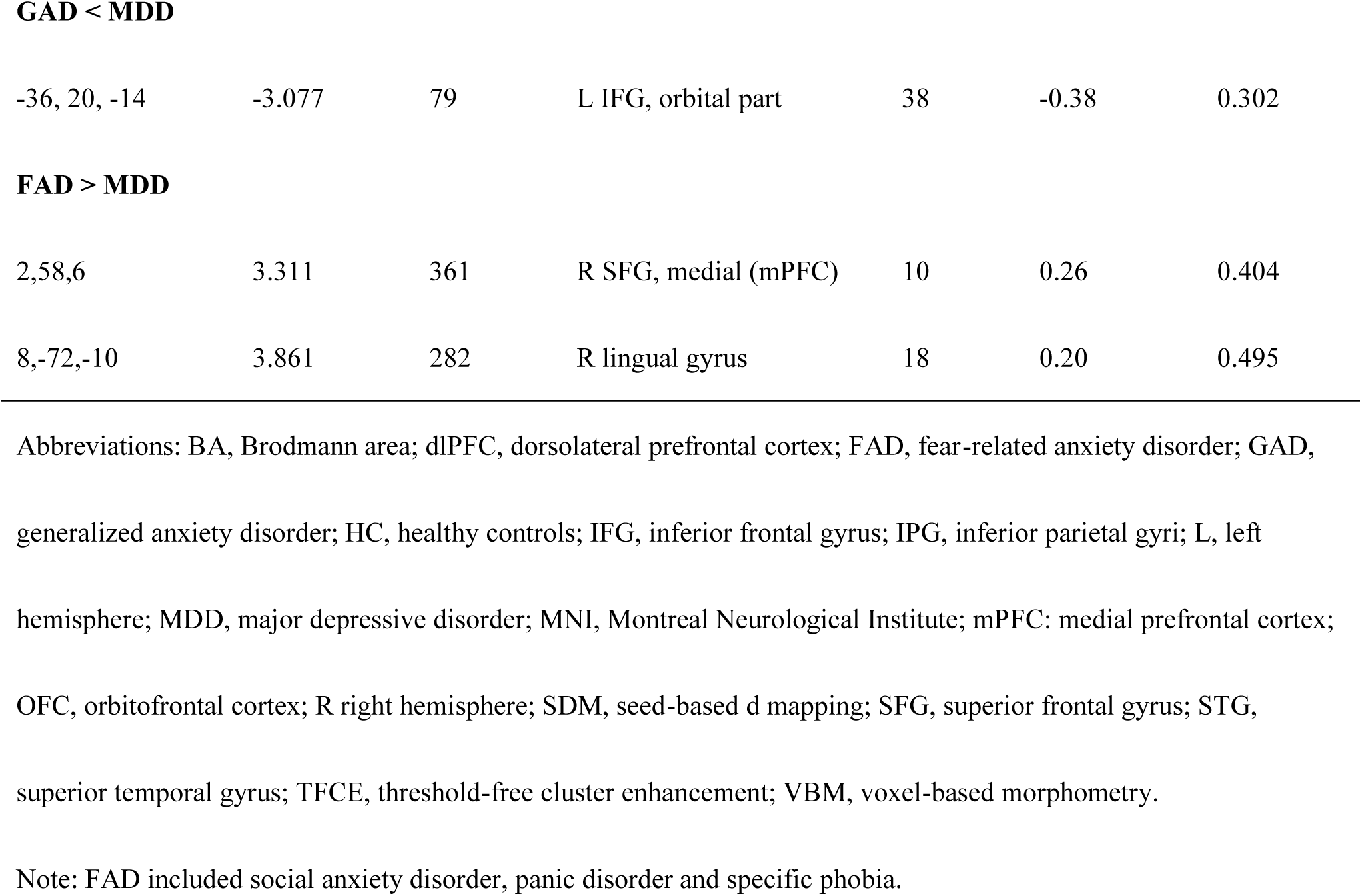
Whole-brain meta-analysis results for VBM studies in GAD, FAD and MDD at threshold TFCE *p* < 0.05.

| MNI coordinates | SDM Z | Voxels | Regions | BA | Egger's bias | Egger's p |
| --- | --- | --- | --- | --- | --- | --- |
| **GAD < HC** |  |  |  |  |  |  |
| -44, -8, 8 | -3.951 | 507 | L Rolandic<br>operculum/insula/STG | 48 | 0.45 | 0.806 |
| -32, 24, -10 | -3.973 | 110 | L IFG, orbital part | 38/47 | -0.41 | 0.809 |
| **FAD vs. HC** |  |  |  |  |  |  |
| *None* |  |  |  |  |  |  |
| **MDD < HC** |  |  |  |  |  |  |
| 2, 36, -10 | -5.012 | 173 | L/R SFG, medial orbital<br>(OFC) | 11 | -0.54 | 0.300 |
| 46, -2, 4 | -4.339 | 115 | R insula | 48 | -0.37 | 0.405 |
| **GAD > FAD** |  |  |  |  |  |  |
| 32, -6, 6 | 1.856 | 153 | R putamen | 48 | 0.50 | 0.654 |
| **GAD < FAD** |  |  |  |  |  |  |
| 2, 54, 16 | -2.102 | 480 | L/R SFG, medial,<br>dorsolateral<br>(mPFC, dlPFC) | 10 | -0.31 | 0.618 |
| -38,-10,10 | -2.981 | 164 | L insula/Rolandic<br>operculum | 48 | -0.37 | 0.513 |
| 52,-48,32 | -2.347 | 136 | R angular gyrus | 48 | -0.35 | 0.530 |
| 44,-36,48 | -2.267 | 41 | R IPG | 2 | -0.34 | 0.552 |
| **GAD < MDD** |  |  |  |  |  |  |
| -36, 20, -14 | -3.077 | 79 | L IFG, orbital part | 38 | -0.38 | 0.302 |
| **FAD > MDD** |  |  |  |  |  |  |
| 2,58,6 | 3.311 | 361 | R SFG, medial (mPFC) | 10 | 0.26 | 0.404 |
| 8,-72,-10 | 3.861 | 282 | R lingual gyrus | 18 | 0.20 | 0.495 |

Abbreviations: BA, Brodmann area; dlPFC, dorsolateral prefrontal cortex; FAD, fear-related anxiety disorder; GAD,
generalized anxiety disorder; HC, healthy controls; IFG, inferior frontal gyrus; IPG, inferior parietal gyri; L, left
hemisphere; MDD, major depressive disorder; MNI, Montreal Neurological Institute; mPFC: medial prefrontal cortex;
OFC, orbitofrontal cortex; R right hemisphere; SDM, seed-based d mapping; SFG, superior frontal gyrus; STG,
superior temporal gyrus; TFCE, threshold-free cluster enhancement; VBM, voxel-based morphometry.
Note: FAD included social anxiety disorder, panic disorder and specific phobia.

**Figure 2.**
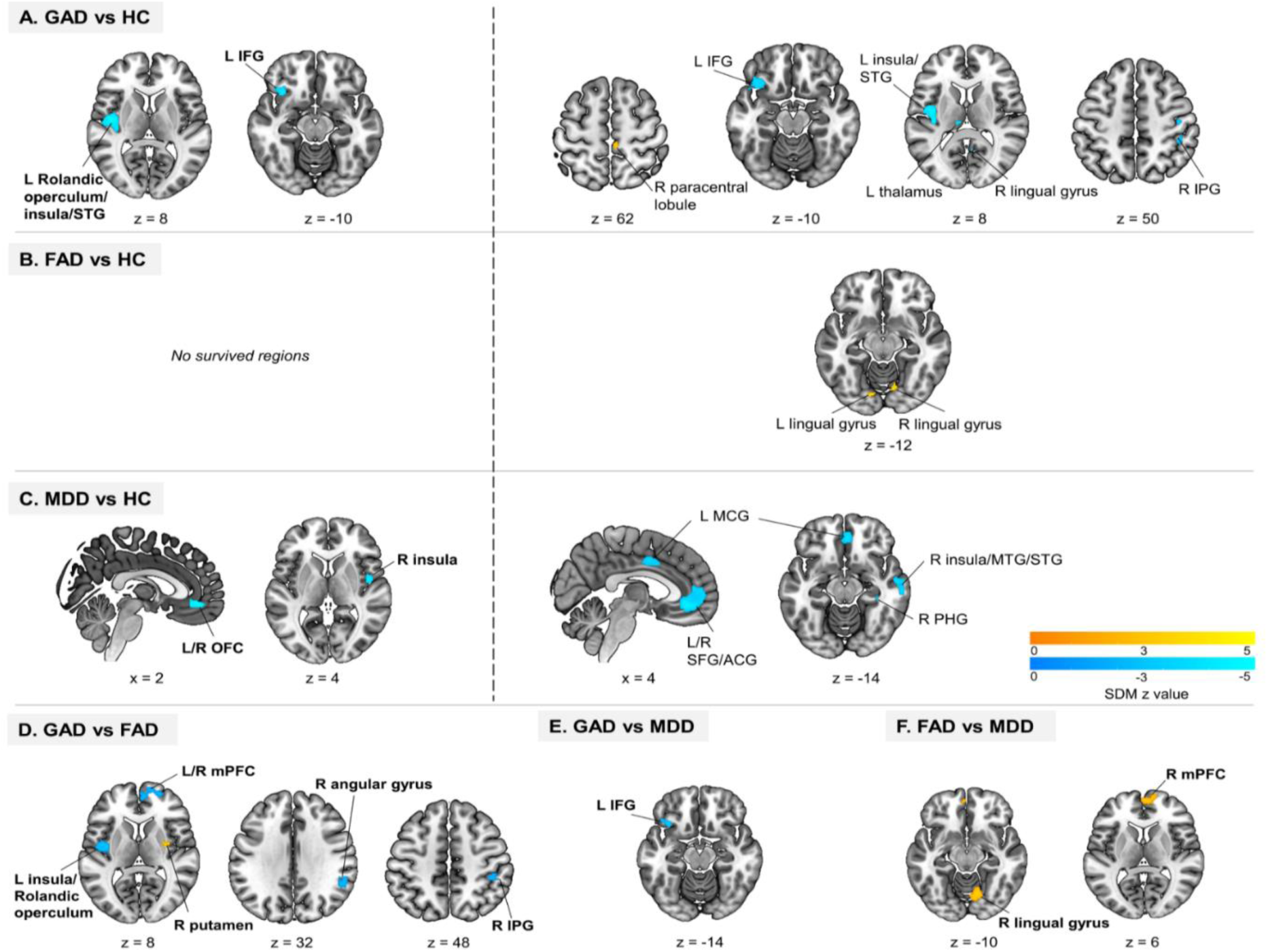
**Results of whole-brain meta-analysis of brain gray matter volume (GMV) in (A) generalized anxiety disorder (GAD) relative to healthy controls (HC), (B) fear-related anxiety disorder (FAD) relative to HC, (C) major depressive disorder (MDD) relative to HC. Left panel shows brain regions significant at *p* < 0.05, TFCE corrected. Right panels show brain regions significant at *p* < 0.0025, uncorrected. (D) GAD (vs HC) in comparison to FAD (vs HC), (E) GAD (vs HC) in comparison to MDD (vs HC), and (F) FAD (vs HC) in comparison to MDD (vs HC). Group comparisons are shown at *p* < 0.05, TFCE corrected.** Abbreviations: ACG, anterior cingulate gyri; IFG, inferior frontal gyrus; IPG, inferior parietal gyrus; L, left; MCG, median cingulate/paracingulate gyri; mPFC, medial prefrontal cortex; MTG, middle temporal gyrus; PHG, parahippocampal gyrus; SFG, superior frontal gyrus; STG, superior temporal gyrus; OFC, orbitofrontal cortex; R, right.

#### 3.2.2 FAD patients versus HC

No significant differences between FAD and HC were found at TFCE-corrected *p* < 0.05 (**Table 2)**. Subgroup meta-analyses in SAD and PD studies yielded no significant GMV alterations. The meta-analysis could not be conducted in SP given the small number of VBM studies (*n* =2). No AG studies were identified.

With a more liberal threshold, we identified increased GMV in the left and right lingual gyrus in FAD (**Table S4, Figure 2B**). Subgroup meta-analyses revealed increased GMV in a wide range of brain regions including the left and right lingual gyrus in SAD, and reduced GMV in the right insula, right STG and left temporal pole/STG in PD (**Table S4**), suggesting neurobiological heterogeneity within FAD groups.

#### 3.2.3 MDD patients versus HC

Patients with MDD relative to HC showed decreased GMV in the OFC and right insula.

No clusters of increased GMV were found (**Table 2, Figure 2C**). With a more liberal threshold, decreased GMV extended into right insula/STG/Rolandic operculum, left and right medial OFC/anterior cingulate cortex (ACC), left and right median cingulate/paracingulate gyri (MCG), and right parahippocampal gyrus (PHG) (**Table S4, Figure 2C**).

#### 3.2.4 Comparison of GMV differences between GAD, FAD and MDD

Covarying for age and sex, there were significant GMV differences between patient groups at *p* < 0.05, TFCE corrected (**Table 2**). GAD was associated with disorder-specific decreased GMV relative to FAD in an mPFC cluster extending to the right dorsolateral PFC, left insula/Rolandic operculum, right angular gyrus and right IPG, whereas FAD had reduced GMV in the right putamen relative to GAD (**Figure 2D**). Disorder-specific reduced GMV was observed in GAD relative to MDD in the left IFG (**Figure 2E**). FAD, relative to MDD, had consistently larger GMV in the right mPFC and lingual gyrus (**Figure 2F**).

#### 3.2.5 GMV Conjunction Analyses

No clusters were identified in all conjunction analyses (i.e., GAD ∩ FAD, GAD ∩ MDD, FAD ∩ MDD, and GAD ∩ FAD∩ MDD) at both *p* < 0.05, TFCE-corrected and *p* < 0.0025, uncorrected.

### 3.3. Meta-regression

To control for potential effects of confounding variables (e.g., age, sex, etc), meta-regression analyses were conducted. Results showed no associations between GMV differences and any confounding variables in GAD and FAD, whereas the higher proportion of medicated MDD was associated with smaller GMV in the left MCG (92 voxels; MNI coordinates: 0, -10, 40; peak *Z* value: -4.323; Brodmann area 23).

### 3.4 Analyses of heterogeneity and publication bias

Extraction of heterogeneity statistics *I*^2^ from the significant clusters indicated low heterogeneity (*I*^2^ <50%). No significant publication bias was revealed by Egger’s test for any significant clusters in GAD, FAD and MDD (*p* > 0.05, **Table 2** and **Table S4**).

## 4. Discussion

The present study capitalized on a robust comparative meta-analytic approach to determine common and disorder-specific neuroanatomical markers in internalizing disorders, specifically in the context of DSM-5 nosology and dimensional models suggesting a differentiation between fear- and anxiety-related AD. After controlling for potential confounders, our meta-analysis demonstrated that, GAD exhibited decreased left insula and IFG volume relative to HC as well as FAD and MDD, respectively. MDD exhibited decreased medial prefrontal volumes relative to HC and FAD yet not GAD, while FAD exhibited increased lingual gyrus volume relative to HC and MDD and decreased putamen volume relative to GAD, yet no GMV alterations in prefrontal regions. No common structural abnormalities were found between the disorders. These findings provide first meta-analytic evidence demonstrating distinct neurobiological alterations in anxiety-related, fear-related, and depressive disorders thus supporting nosologically distinguishable entities.

Initial meta-analyses within each disorder group resembled previous meta-analytic findings in GAD (22) and MDD (39). Previous meta-analyses employed less stringent approaches and commonly reported more widespread and regionally unspecific neuroanatomical alterations. The present results, however, suggested highly robust and regional-specific structural alterations at a strict inference level in GAD or MDD, respectively. In contrast to our hypothesis, no convergent GMV alterations were observed in FAD, which may result from the heterogeneity of FAD and is consistent with a recent meta-analysis indicating no shared GMV alterations between SAD and PD (24). A more lenient threshold gave rise to increased GMV in the bilateral lingual gyrus and subgroup analyses suggested that increases in the lingual gyrus were mainly driven by SAD. Although this resembles prior original VBM studies reporting greater GMV in the lingual gyrus in SAD (40), the current results may reflect a neuroanatomical heterogeneity of FAD originating from the overarching ‘fear’ pathology meta-factor (6, 8).

The comparative approach allowed us to further dissect disorder-specific and transdiagnostic alterations. Robust volumetric reductions in the left Rolandic operculum/insula/STG and IFG differentiated GAD from HC as well as FAD and MDD, respectively, indicating a regional- and GAD-specific neuroanatomical marker which is consistent with previous systematic reviews and meta-analyses describing insula and PFC impairments in GAD (22, 41). Notably, our study provides the first meta-analytic evidence demonstrating reduced insula volume in GAD relative to FAD, which aligns with a recent neurofunctional meta-analysis showing a GAD-specific hypoactivation of the bilateral insula across cognitive and emotion domains in contrast to other AD (36). The insula/Rolandic operculum are implicated in interoceptive, salience and emotion processing (42). Resting-state fMRI and lesion studies suggested that deficits in Rolandic operculum are associated with GAD and high anxiety and perceived stress which characterizes GAD (43, 44). The left IFG has been consistently reported in previous meta-analyses examining brain structural markers of both, GAD (22) and mixed AD (27, 28). The IFG plays a critical role in GAD with reduced GMV in geriatric GAD patients (45), disorder-specific hypoactivation during emotion regulation relative to PD (15) or MDD (19) and dysfunctional connectivity in GAD (46). The left IFG is specifically critical for top-down regulatory control (47). Therefore, disorder-specific reduced GMV in the left insula/Rolandic operculum and IFG may reflect altered emotion processing of potential threats and inhibitory response to worrisome thoughts in GAD.

Notably, both GAD and MDD exhibited smaller GMV in the mPFC in comparison to FAD. fMRI studies have shown common neurofunctional alterations in the mPFC and ACC in cognitive or emotional processing in both GAD and MDD (17–19). Previous meta-analyses have identified altered volume in mPFC in GAD (22), MDD (26), and individuals with high neuroticism (48), a pathological meta-factor associated with GAD and MDD which share symptomatic (e.g., negative affect, worry) (4) and genetic etiologies (5). The mPFC is a key node in the default mode network (DMN), which is engaged in self-referential processing and emotion regulation including distress tolerance (49, 50). Behavioral studies suggest that individuals with a distress-misery disorder history (e.g., GAD and MDD) perceive less capacity to tolerate negative affect than fear disorders (e.g., SAD,PD and SP) (51). Thus, the mPFC deficit in GAD and MDD relative to FAD may reflect a neurobiological marker for common anxious-misery disorders characterized by inappropriate self-referential thinking and uncontrollable worry and distress.

Evidence for robust and disorder-specific neuroanatomical markers in FAD was less consistent, yet some evidence for increased lingual gyrus volume relative to both, HC and MDD, as well as decreased putamen volume in comparison to GAD was found. The lingual gyrus is suggested to be associated with panic symptoms in MDD (52) and involved in abnormal sensory gating of emotional stimuli such as facial expressions in SAD and PD (40, 52). Consistent with our findings, previous studies reported greater GMV and cortical thickness in the lingual gyrus in SAD relative HC (40) and MDD (53). The smaller GMV in right putamen in FAD relative to GAD aligns with previous meta-analytic findings demonstrating decreased putamen GMV in FAD relative to HC and obsessive-compulsive disorders (OCD) (23, 24, 54), as well as with original studies reporting larger putamen volume in GAD (55). The putamen is a part of the dorsal striatum and is related to anxiety disorders and symptoms (56, 57). For example, disorder-specific heightened putamen activation during incentive anticipation was found in SAD relative to HC and GAD (56) and putamen volume has been found to be negatively correlated with the severity of PD symptoms (58). Positron emission tomography (PET) studies have shown that SAD and PD is associated with compromised serotonergic (5-HT) neurotransmission in several brain areas including the putamen (59). Decreasing the function of the 5HT system has been reported to exacerbate psychological and physiological response to stressors in fear disorders (e.g., PD, SAD), but not in anxiety disorders (e.g., GAD, OCD) (60). Together, our findings suggest that putamen volume differences may represent a dissociable neuroanatomical deficit in fear-versus anxiety-related AD, and in the context of previous studies decreased putamen GMV in FAD may be associated with serotonin dysfunction.

The results in MDD replicated previous meta-analyses reporting GMV reductions in bilateral OFC and right insula (26). Functional and structural deficits of the OFC have been repeatedly reported in MDD and are associated with the severity of rumination (61) and depressive symptom (62) in MDD, which may underlie the cognitive, mood and social impairments. The insula has been found to play an important role in the pathophysiology of MDD (63, 64) and predict treatment response in MDD (65). Our study specifically identified reduced GMV in right mid-posterior insula in MDD, consistent with fMRI studies showing that interoceptive abnormalities in MDD are associated with bilateral mid-posterior insula dysfunction (66). The mid-posterior insula has been associated with interoception, somatosensory, and pain (67). Reduced GMV in mid-posterior insula has also been described in other mental disorders such as post-traumatic stress disorder, schizophrenia and anorexia nervosa (64), suggesting that GMV reduction in this region may characterize mental disorders with dysfunctions in interoceptive processing. However, these previous studies and the current study on GAD revealed alterations in left mid-posterior insula, whereas our study points towards right mid-posterior insula in MDD. Therefore, although insula may be a potential biomarker for anxious-misery disorders, the dissociation of the left and right mid-posterior insula in GAD and MDD needs to be disentangled in future studies.

In contrast to our hypothesis, the conjunction analyses did not yield common neuroanatomical alterations in the disorders. Previous meta-analyses revealed transdiagnostic neural markers of psychopathology pointing towards common deficits in ACC/PFC and insula both structurally (31) and functionally (32). However, these meta-analyses included both psychotic (e.g., schizophrenia) and non-psychotic disorders (e.g., substance use disorder, and OCD) and did not specifically aim at determining transdiagnostic alterations within internalizing disorders. Methodologically, the identification of transdiagnostic alterations in previous meta-analyses may result from the utilization of less stringent correction approaches (68). In contrast, the present meta-analysis suggests separable rather than unspecific neuroanatomical abnormalities in fear, anxiety and depressive disorders.

Several limitations should be considered. First, the number of studies in GAD is relatively small. Although it meets the recommended number of studies for use of SDM and one original map is included, the statistical power for GAD is limited compared to FAD and MDD. Second, the unbalanced number of studies in FAD subtypes and the two original maps included for SAD may bias the results towards specific subtypes of FAD such as SAD.

Finally, conceptually we only included GAD as a representative category of anxiety-related anxiety disorder. Other anxiety-related disorders such as separation anxiety disorder were not included because there are no clear neural components for these categories.

## 5. Conclusion

Summarizing, our study is the first to comprehensively investigate common and distinct GMV alterations in anxiety- and fear-related anxiety disorders, as well as depressive disorders, which bridges a gap between current psychiatric nosology and neurobiological findings. In line with previous definitions, we found distinct neuroanatomical substrates underlying the pathophysiology of GAD, FAD and MDD with dissociated prefrontal and insula deficits in GAD and MDD characterized by anxious-misery, and striatum and threat detection deficits in FAD characterized by fear. The disorder-specific biomarkers could serve as future therapeutic targets.

## Data Availability

Coordinates and t-value files are available at https://osf.io/46uc2/. Unthresholded whole-brain maps are provided at https://neurovault.org/collections/11343/.

https://osf.io/46uc2/

## Acknowledgments and disclosures

This work was supported by the National Key Research and Development Program of China (Grant No. 2018YFA0701400). We thank the authors who provided whole-brain *t*-maps of their original studies.

Dr. Becker, Dr. Zhang, Dr. Klugah-Brown, Ms. Liu and Ms. Zhang report no financial relationships with commercial interests.

## Notes

### Competing Interest Statement

The authors have declared no competing interest.

### Clinical Protocols

https://osf.io/es2vm

