## Supplementary for "Distinct neurostructural signatures of anxiety-, fear-related and depressive disorders: a comparative voxel-based meta-analysis"

### Supplement Methods

#### Literature search and inclusion

Database search keywords for Generalized Anxiety Disorder (GAD) included: (“generalized anxiety disorder”, OR (“GAD”)) AND (“voxel-based morphometry”, OR (“VBM”), OR (“gray matter”)).

Database search keywords for Fear-related Anxiety Disorders (FAD) included: (“social anxiety disorder”, OR (“social phobia”), OR (“SAD”), OR (“specific phobia”), OR (“panic disorder”), OR (“Phobia”), OR (“agoraphobia”)) AND (“voxel-based morphometry”, OR (“VBM”), OR (“gray matter”)).

Database search keywords for Major Depressive Disorder (MDD) included: “Depression” AND (“voxel-based morphometry”, OR (“VBM”), OR (“gray matter”)). Broad search terms were used to prevent missing any relevant studies. Additionally, we inspected reference lists of relevant reviews and original studies to identify further eligible articles.

Key inclusion criteria were (1) neuroimaging case control design studies comparing brain structure between a group of patients with GAD, FAD or MDD with healthy controls (HC), and (2) whole-brain VBM between-group comparisons (including null findings) reported. Studies were excluded if they (1) were non-empirical studies (e.g., review, meta-analysis, meeting abstract); (2) were not published in English; (3) did not report direct comparison between a patient and healthy control (HC) group; (4) did not report whole-brain results (i.e., only ROI results); (5) did not provide sufficient data (e.g., peak coordinates, etc.) even after contacting the authors of the original study. In case the same HC reference group was employed for several subgroup comparisons, only the combined result was included in the meta-analysis.

#### Meta-analysis

*SDM procedures*

For each article group, raw statistical maps or coordinates from cluster peaks including their effective sizes (t-values, representing the GMV differences between patients and controls) and sample sizes were entered into the SDM-PSI to be preprocessed using a 20 mm full width half maximum (FWHM) anisotropic Gaussian kernel and 2 mm voxel size. In this step, SDM converts *t*-values into Hedge's g effect sizes and their variances using evaluated algorithms (1) in case raw maps for the original study are not available. SDM conducts multiple imputation to estimate the lower and upper bounds of possible effect sizes for each study using an anisotropic Gaussian kernel. Then in the mean analysis, SDM-PSI conducts unbiased maximum likelihood estimation (MLE) of effect sizes and associated standard errors by imputing the effect size maps of individual studies for each group based on MetaNSUE algorithms (2, 3). To prevent a single study or few studies from driving the results, SDM uses a leave one out jackknife procedure. The number of imputations for mean analysis was set to 50 by default. Finally, SDM-PSI allows a Freedman-Lane-based permutation test at the subject level to perform voxel-wise tests of statistical significance while controlling familywise error rate (FWER) using threshold-free cluster enhancement (TFCE) (4). Details of these procedures are provided in the corresponding methodological publications (5, 6) and the SDM-PSI reference manual (https://www. sdmproject.com/manual/).

*Transdiagnostic meta-analysis*

A transdiagnostic meta-analysis of VBM studies across GAD (9 studies, 226 patients), FAD (23 studies, 996 patients) and MDD (46 studies, 2575 patients) was conducted and thresholded at TFCE-based FWE corrected $p < 0.05$.

Partly resembling the results of a previous transdiagnostic meta-analysis including schizophrenia, bipolar disorder, major depression, substance use disorder and a pooled group of anxiety disorders (7), we observed decreased right insular cortex volume in the overarching pooled meta-analysis (**Figure. S1**). However, the number of MDD studies was at least two times larger than the number of GAD or FAD studies and the imbalance may strongly bias the results in favor of the MDD-related alterations (see also results in the main text demonstrating reduced right mid-insula volumes in MDD but not FAD or GAD, **Figure 2A, B, C**), while the conjunction approach in our main analyses would only identify regions showing convergent alterations across the disorder categories.

#### Supplement Results

**Table S1. Demographic and clinical characteristics of the 9 GAD VBM datasets included in the meta-analysis.**

| Study | Number (female) | | Age (years) | | Duration (years) | Medication | Comorbidity | Scanner/ FWHM (mm) | *p* value | Summary findings |
|---|---|---|---|---|---|---|---|---|---|---|
| | GAD | HC | GAD | HC | | | | | | |
| Chen et al. 2020 * | 72 (41) | 57 (30) | 39.04 | 40.91 | 4.56 | Medication load index: 1.67 ± 0.69 | 0 | 3T/8 | $p < 0.05$ (GRF) | GAD < HC: L/R sgACC/vmPFC, L ITG, R Insula, R dmPFC |
| Hilbert et al. 2015 | 19 (16) | 24 (17) | 33.47 | 32.25 | NA | Drug naïve | 12 MDD, 2 dysthymia, 13 other anxiety disorders | 3T/8 | $p < 0.05$ (FWE) | GAD > HC: R Striatum |

| | | | | | | | | | | |
|---|---|---|---|---|---|---|---|---|---|---|
| Kim et al. 2018 | 16 (6) | 16 (6) | 33.00 | 32.50 | 5.60 | Medicated (*n* = 10) | NA | 3T/6 | $p < 0.001$ (uncorr) | GAD < HC: R ACC, L Precuneus, L dlPFC, L OFG, L SOG, L Insula, L STG, L mPFC |
| Liao et al. 2014 | 26 (13) | 25 (12) | 16.85 | 16.72 | NA | Drug naïve | 0 | 3T/8 | $p < 0.05$ (FWE) | GAD > HC: R Putamen |
| Ma et al. 2019 | 21 (9) | 20 (10) | 34.92 | 35.96 | 2.39 | Drug-free (> 6 month) | 0 | 3T/6 | $p < 0.05$ (AlphaSim) | GAD < HC: R Precentral gyrus, R SFG |
| Makovac et al. 2016 | 19 (16) | 19 (16) | 30.00 | 29.20 | 16.78 | Medicated (*n* = 2) | 0 | 1.5T/8 | $p < 0.05$ (FWE) | GAD < HC: R Supramarginal gyrus/postcentral gyrus, L Postcentral gyrus, R Precentral |

| | | | | | | | | | | |
|---|---|---|---|---|---|---|---|---|---|---|
| | | | | | | | | | | gyrus, L Supramarginal gyrus |
| Moon et al. 2014 | 22 (9) | 22 (9) | 37.00 | 33.40 | 4.90 | NA | NA | 3T/8 | $p < 0.001$ (uncorr) | GAD < HC: L Midbrain, L Thalamus, L Hippocampus, L Insula, L STG |
| Schienle et al., 2011 | 16 (16) | 15 (15) | 22.90 | 23.70 | 3.10 | Drug naïve | 0 | 3T/12 | $p < 0.05$ (FWE) | -- |
| Strawn et al., 2013 | 15 (8) | 28 (17) | 13.00 | 13.00 | NA | Drug naïve | 6 ADHD, 6 other anxiety disorders | 4T/8 | $p < 0.001$ (uncorr) | GAD > HC: R precentral gyrus, R Precuneus; GAD < HC: R PCC, L OFG |

Abbreviations: ACC, anterior cingulate cortex; dlPFC, dorsolateral prefrontal cortex; dmPFC, dorsomedial prefrontal cortex; FWE, family-wise error; FWHM, full width at half maximum; GAD, generalized anxiety disorder; GRF, Gaussian random field; HC, healthy controls; ITG, inferior temporal gyrus; L, left hemisphere; mPFC, medial prefrontal cortex; NA, not available; OFG, orbitofrontal gyrus; PCC, posterior cingulate cortex; R right hemisphere; SFG, superior frontal gyrus; sgACC, subgenual anterior cingulate cortex; SOG, superior occipital gyrus; STG, superior temporal gyrus; uncorr, uncorrected; VBM, voxel-based morphometry; vmPFC ventromedial prefrontal cortex.

* Studies that provided original whole-brain *t*-maps.

**Table S2. Demographic and clinical characteristics of the 32 FAD VBM datasets included in the meta-analysis.**

| Study | Disorder type | Number (female) | | Age (years) | | Duration (years) | Medication | Comorbidity | Scanner/ FWHM (mm) | *p* value | Summary findings |
|---|---|---|---|---|---|---|---|---|---|---|---|
| | | Patients | HC | Patients | HC | | | | | | |
| Bas-Hoogendam et al. 2017 * | SAD | 174 (102) | 213 (106) | 30.60 | 32.40 | 15.80 | Medicated (n = 24) | 8 MDD, 2 MDD + PD, 10 GAD, 3 GAD + SP, 2 GAD + PD, 3PD, 6 SP, 26 unknown | 3T/7.5 mm | $p < 0.05$ (FWE) | -- |
| Cheng et al. 2015 | SAD | 20 (7) | 30 (9) | 23.30 | 26.20 | 3.99 | Drug naïve | 0 | 3T/8 mm | $p < 0.05$ (FWE) | -- |
| Frick et al. 2014 | SAD | 48 (24) | 29 (16) | 33.80 | 23.70 | NA | Drug naïve | 10 GAD; 7 SP; 3 MDD; 2 PD | NA/8 mm | $p < 0.05$ (corr) | SAD > HC: L/R Lingual gyrus, L |

| | | | | | | | | | | | |
|---|---|---|---|---|---|---|---|---|---|---|---|
| | | | | | | | | | | | FFG, R Secondary occipital cortex |
| Irle et al. 2014 | SAD | 67 (35) | 64 (31) | 31.00 | 32.00 | 15.00 | Medicated (n = 6) | 16 MDD; 7 SP; 5 PD; 1 GAD | 3T/8 mm | *p* < 0.001 (uncorr) | -- |
| Kawaguchi et al. 2016 | SAD | 13 (8) | 18 (0) | 36.20 | 33.80 | 23.30 | NA | 4 MDD; 1 PD | 3T/8 mm | *p* < 0.05 (FWE) | -- |
| Liao et al. 2011 | SAD | 18 (6) | 18 (5) | 22.70 | 21.90 | 4.10 | Drug naïve | 0 | 3T/8 mm | *p* < 0.05 (AlphaSim) | SAD > HC: R mPFC; SAD < HC: L PHG, R ITG |
| Meng et al. 2013 | SAD | 20 (6) | 19 (6) | 21.80 | 21.60 | 4.21 | Drug naïve | 0 | 3T/12 mm | *p* < 0.05 (AlphaSim) | SAD < HC: L/R Thalamus, R |

| | | | | | | | | | | | |
|---|---|---|---|---|---|---|---|---|---|---|---|
| Talati et al. 2013 | SAD | 33 (9) | 37 (19) | 31.50 | 31.40 | NA | Medicated (n = 9) | 11 MDD;5 GAD; 4 SP, 1 OCD, 1 DUD, 2 AUD | 1.5T/8 mm | $p < 0.05$ (nonstationary corr) | Amygdala, R Precuneus SAD > HC: R supramarginal and angular cortices; SAD < HC: L temporal pole |
| Tükel et al. 2015 * | SAD | 27 (15) | 27 (15) | 27.70 | 27.70 | 13.80 | Drug naïve | 0 | 1.5T/8 mm | $p < 0.05$ (AlphaSim) | SAD > HC: L Superior parietal and precuneus, R middle and inferior temporal areas, R FFG |

| | | | | | | | | | | | |
|---|---|---|---|---|---|---|---|---|---|---|---|
| Zhao et al. 2017 | SAD | 24 (9) | 41 (15) | 24.50 | 27.10 | 7.60 | Drug naïve | 0 | 3T/8 mm | *p* < 0.001 (FDR) | SAD < HC: L/R Putamen, L/R OFC, L/R Thalamus |
| Asami et al. 2009 | PD | 24 (15) | 24 (15) | 37.03 | 37.01 | 3.90 | Medicated (n = 36) | 13 AG, 3 MDD, 1 dysthymia | 1.5T/12 mm | *p* < 0.05 (FDR) | PD < HC: L/R dmPFC, R vmPFC, R Amygdala, R ACC, L/R STG, L/R Insula, L/R Lateral occipitotemporal gyrus, L cerebellar vermis |

| | | | | | | | | | | | |
|---|---|---|---|---|---|---|---|---|---|---|---|
| Kunas et al. 2020 | PD | 143 (89) | 178 (101) | 33.65 | 31.63 | NA | Drug naïve | 51 MDD | 3T/8 mm | $p <$ 0.001 (uncorr) | PD < HC: L MTG |
| Lai & Wu, 2012 | PD | 30 (19) | 21 (11) | 47.03 | 41.14 | NA | Drug naïve | 13 AG | 3T/7.5 mm | $p <$ 0.005 (FWE) | PD < HC: L OFC, L IFG, R Insula, L STG |
| Lai & Wu, 2015 | PD | 53 (28) | 54 (29) | 43.28 | 40.38 | 5.35 | Drug naïve | 0 | 3T/7.5 mm | $p < 0.05$ (FWE) | PD < HC: R IFG R Insula |
| Massana et al. 2003 | PD | 18 (11) | 18 (10) | 36.80 | 36.70 | NA | Drug naïve | 15 AG | 1.5T/12 mm | $p < 0.05$ (corr) | PD < HC: L PHG |
| Na et al. 2013 | PD | 22 (9) | 22 (11) | 40.18 | 40.18 | NA | Drug naïve | 12 AG | 3T/8 mm | $p < 0.05$ (FWE) | PD < HC: L/R SOG, cuneus |
| Protopopescu et al. 2006 | PD | 10 (6) | 23 (11) | 33.50 | 28.70 | NA | Medicated (n = 1) | 2 AG | 3T/12 mm | $p < 0.05$ (GRF) | PD > HC: Brainstem |

| | | | | | | | | | | | |
|---|---|---|---|---|---|---|---|---|---|---|---|
| Sobanski et al. 2010 | PD | 17 (9) | 17 (9) | 34.90 | 33.10 | 7.90 | NA | 16 AG, 1 SAD, 1 GAD, 2 AD, 2 PPD | 1.5T/12 mm | *p* < 0.05 (FWE) | PD < HC: R MTG, R OFC |
| Talati et al. 2013 | PD | 16 (13) | 20 (9) | 31.80 | 31.40 | 13.40 | Medicated (n = 9) | 3 MDD, 2 GAD,5 SP, 2 OCD,4 DUD,4 AUD | 1.5T/8 mm | *p* < 0.05 (corr) | PD > HC: L/R Cuneus, lingual; PD < HC: R precentral, postcentral, R middle cingulate |
| Uchida et al. 2008 | PD | 19 (16) | 20 (16) | 37.05 | 36.45 | 8.34 | Medicated (n = 15) | 14 AG, 3 MDD, 2 dysthymia | 1.5T/12 mm | *p* < 0.001 (uncorr) | PD > HC: L Insula and STG, L Midbrain and pons; PD < HC: R ACC |

| | | | | | | | | | | | |
|---|---|---|---|---|---|---|---|---|---|---|---|
| Yoo et al. 2005 | PD | 18 (9) | 18 (7) | 33.30 | 32.00 | 3.60 | Medicated (n = 10) | 0 | 3T/8 mm | $p < 0.05$ (corr) | PD < HC: L/R Putamen, R Precuneus gyrus, R ITG, L STG, L SFG |
| Hilbert et al. 2015 | SP | 59 (45) | 37 (28) | 23.95 | 22.76 | NA | Drug naïve | 0 | 1.5T/12 mm | $p < 0.05$ (FWE) | Specific phobia > HC: R ACC, R Calcarine sulcus, R FFG, L medial OFC, L Precuneus, R Vermis |
| Schienlei et al. 2013 | SP | 45 (25) | 41 (23) | 30.39 | 29.24 | 16.69 | Drug naïve | 0 | 3T/8 mm | $p < 0.001$ (uncorr) | -- |

Abbreviations: ACC, anterior cingulate cortex; AD, adjustment disorder with mixed disturbance of emotions and conduct; AG, agoraphobia; AUD, alcohol use disorder; corr, corrected; dlPFC, dorsolateral prefrontal cortex; dmPFC, dorsomedial prefrontal cortex; DUD, drug use disorder; FAD, fear-related anxiety disorder; FDR, false discovery rate; FFG, fusiform gyrus; FWE, family-wise error; FWHM, full width at half maximum; GRF, Gaussian random field; HC, healthy controls; IFG, inferior frontal gyrus; IOG, inferior occipital gyrus; IPL, inferior parietal lobule; ITG, inferior temporal gyrus; L, left hemisphere; LOC, lateral occipital cortex; MCC, medial cingulate cortex ; MDD, major depressive disorder; MFG, medial frontal gyrus; MOG, middle occipital gyrus; mPFC, medial prefrontal cortex; MTG, middle temporal gyrus; NA, not available; OCD, obsessive-compulsive disorder; OFC, orbitofrontal cortex; PCC, posterior cingulate cortex; PCG, paracingulate gyrus; PD, panic disorder; PHG, parahippocampal gyrus; PPD, paranoid personality disorder; PTSD, post-traumatic stress disorder; R right hemisphere; SAD, social anxiety disorder; SFG, superior frontal gyrus; SMA, supplementary motor area; SOG, superior occipital gyrus; SP, specific phobia; STG, superior temporal gyrus; TOFC, temporal-occipital fusiform cortex; uncorr, uncorrected; VBM, voxel-based morphometry; vmPFC ventromedial prefrontal cortex.

* Studies that provided original whole-brain *t*-maps.

**Table S3. Demographic and clinical characteristics of the 46 MDD VBM datasets included in the meta-analysis.**

| Study | Number (female) | | Age (years) | | Duration (years) | Medication | Comorbidity | Scanner/ FWHM (mm) | *p* value | Summary findings |
|---|---|---|---|---|---|---|---|---|---|---|
| | MDD | HC | MDD | HC | | | | | | |
| Abe et al. 2010 | 21 (10) | 42 (20) | 48.10 | 48.00 | 6.00 | Medicated (n = 19) | 0 | 1.5T/6 mm | $p < 0.05$ (FDR) | MDD < HC: R PHG, L/R Middle frontal gyrus, L IOG, L SMA, R STG, L Parietal lobe, L/R ACC |
| Ahn et al. 2016 | 34 (29) | 26 (19) | 32.43 | 31.40 | NA | NA | NA | 3T/8 mm | $p < 0.05$ (AlphaSim) | MDD > HC: L Postcentral gyrus, R Postcentral gyrus, L/R Parieto-occipital cortex, L/R Putamen, R Thalamus, L/R Hippocampus, L/R Cerebellum; |

| | | | | | | | | | | |
|---|---|---|---|---|---|---|---|---|---|---|
| | | | | | | | | | | MDD < HC: L/R OFC, R dmPFC, R dorsal ACC, L/R MOG, L Cuneus |
| Amico et al. 2011 | 33 (14) | 64 (28) | 32.00 | 30.40 | 3.40 | Medicated (n = 27) | 0 | 1.5T/8 mm | $p < 0.05$ (FWE) | -- |
| Arnone et al. 2013 | 39 (27) | 66 (46) | 36.30 | 32.10 | 14.30 | Drug naïve | 0 | 1.5T/8 mm | $p < 0.05$ (FWE) | MDD < HC: L/R Hippocampus/PHG, L/R FFG/ITG, L/R Ventral striatum |
| Bergouignan et al. 2009 | 20 (17) | 21 (14) | 33.16 | 28.21 | 8.45 | Medicated (n = 20) | 0 | 1.5T/8 mm | $p < 0.05$ (FDR) | MDD < HC: R Cingulate gyrus, R MTG, R Posterior lobe, R SPL, R PHG, L Inferior semi-lunar lobule |

| | | | | | | | | | | |
|---|---|---|---|---|---|---|---|---|---|---|
| Biedermann et al., 2015 | 46 (34) | 35 (22) | 50.80 | 46.40 | NA | Medicated (n = 24) | NA | 1.5T/8 mm | $p < 0.05$ (FWE) | -- |
| Cai et al., 2015 | 23 (10) | 23 (10) | 30.00 | 30.00 | 4.35 | NA | 0 | 3T/8 mm | $p < 0.001$ (uncorr) | MDD < HC: IFG |
| Chaney et al. 2014 | 37 (21) | 46 (28) | 40.22 | 36.61 | 9.65 | Medicated (n = 24) | 0 | 3T/10 mm | $p < 0.05$ (FWE) | -- |
| Chen et al. 2016 * | 27 (14) | 28 (14) | 33.00 | 33.00 | 6.58 | Drug naïve | 0 | 3T/6 mm | $p < 0.05$ (AlphaSim) | MDD > HC: R Postcentral gyrus |
| Chen et al. 2020 | 22 (18) | 22 (18) | 28.70 | 27.40 | 4.40 | Medicated (n = 17) | 0 | 3T/8 mm | $p < 0.001$ (uncorr) | MDD < HC: R Occipital fusiform gyrus, L Postcentral gyrus |
| Dannlowski et al. 2015 | 171 (105) | 512 (289) | 38.60 | 33.30 | NA | Medicated (n = 171) | 0 | 3T/8 mm | $p < 0.001$ (Monte Carlo) | MDD < HC: R PHG/HC/amygdala, R Insula/putamen, IFG, L |

| | | | | | | | | | | |
|---|---|---|---|---|---|---|---|---|---|---|
| | | | | | | | | | | Lingual gyrus, FFG, Cerebellum, IOG, L/R MCC, L STG/MTG, L/R Thalamus |
| Fang et al. 2015 | 20 (8) | 18 (8) | 59.20 | 59.10 | 3.60 | NA | NA | 1.5T/8 mm | $p < 0.05$ (corrected) | MDD < HC: L/R ITG, L IFG, L Precuneus, R MCC, R FFG, R Cuneus |
| Förster et al. 2020 | 63 (33) | 46 (22) | 42.43 | 45.35 | NA | NA | 19 anxiety disorders | 3T/8 mm | $p < 0.05$ (FWE) | -- |
| Grieve et al. 2013 | 102 (54) | 34 (16) | 33.80 | 31.50 | 11.30 | NA | 0 | 3T/8 mm | $p < 0.05$ (FDR) | MDD < HC: R Rectal gyrus, R IFG, L FFG, R ITG, L MTG, |
| Hagan et al. 2015 | 109 (81) | 36 (26) | 15.56 | 15.65 | NA | Medicated (n = 37) | 0 | 3T/7 mm | $p < 0.05$ (FWE) | -- |
| Inkster et al. 2011 | 145 (94) | 183 (110) | 49.45 | 48.00 | 14.30 | NA | 0 | 1.5T/10 mm | $p < 0.05$ (FWE) | MDD < HC: R SPL |

| | | | | | | | | | | |
|---|---|---|---|---|---|---|---|---|---|---|
| Kandilarova et al. 2019 | 39 (29) | 42 (29) | 47.70 | 42.60 | 10.80 | Medicated (n = 37) | 0 | 3T/8 mm | $p < 0.05$ (FDR) | MDD < HC: L MFG/ACC |
| Klauser et al. 2015 | 56 (40) | 33 (21) | 34.02 | 34.71 | 10.29 | Medicated (n = 33) | 0 | 1.5T/10 mm | $p < 0.05$ (permutation) | MDD < HC: L/R vmPFC |
| Lee et al. 2011 | 47 (42) | 51 (45) | 46.00 | 45.70 | 3.89 | Medicated (n = 27) | 0 | 1.5T/10 mm | $p < 0.05$ (FDR) | MDD < HC: L/R Midbrain, L/R sgACC, L/R Thalamus, L Short insular gyrus, L/R Nucleus accumbens, L/R Amygdala, L/R Hippocampus, L/R FFG, L Lingual gyrus, L/R MTG, R STG, L/R Cerebellum |

| | | | | | | | | | | |
|---|---|---|---|---|---|---|---|---|---|---|
| Lee et al. 2020 | 20 (20) | 21 (21) | 42.50 | 42.30 | 9.50 | Medicated (n = 20) | 0 | 1.5T/10 mm | *p* < 0.05 (FDR) | MDD < HC: L/R Middle frontal gyrus, L/R Rectal gyrus, L/R Short and long insula gyrus, L/R ACC, MCC, L/R PCC, L/R Thalamus, L/R Hypothalamus, L/R Amygdala, L/R Hippocampus, L/R PHG, L Lingual gyrus, L/R STG, MTG, ITG |
| Leung et al. 2009 | 17 (17) | 17 (17) | 45.50 | 45.80 | 7.00 | Medicated (n = 17) | NA | 1.5T/12 mm | *p* < 0.001 (uncorr) | MDD < HC: R ACC, R STG, R SFG, L/R MFG, R FFG, R IFG, L/R Precentral gyrus, L MCC, L Insula, L Angular |

| | | | | | | | | | | |
|---|---|---|---|---|---|---|---|---|---|---|
| | | | | | | | | | | gyrus, L Precuneus, L MTG, L MCC, PCC |
| Lu et al. 2018 | 76 (44) | 86 (43) | 34.27 | 33.35 | 3.74 | Drug naïve | 0 | 3T/8 mm | *p* < 0.05 (GRF) | MDD > HC: L L Caudate, R ITG, L Cerebellum, L Lingual gyrus; MDD < HC: L/R PHG/Hippocampus, R MTG |
| Lu et al. 2019 | 30 (13) | 48 (30) | 23.98 | 21.50 | 2.56 | Drug naïve | 0 | 3T/8 mm | *p* < 0.05 (GRF) | MDD > HC: R MOG; MDD < HC: L SPL |
| Mak et al. 2009 | 17 (17) | 17 (17) | 45.50 | 45.80 | NA | Medicated (n = 17) | 0 | 1.5T/8 mm | *p* < 0.001 (uncorr) | MDD < HC: R ACC, R SMA, R Precentral gyrus, R Temporal pole, L Angular gyrus, L Precuneus |

| | | | | | | | | | | |
|---|---|---|---|---|---|---|---|---|---|---|
| Modinos et al. 2014 | 23 (20) | 46 (14) | 44.60 | 25.30 | NA | NA | NA | 1.5T/8 mm | *p* < 0.05 (FWE) | MDD < HC: L/R Orbital gyrus, R MFG, L ACC, L/R IPL, R MTG, L MFG |
| Nakano et al. 2014 | 36 (22) | 54 (27) | 49.00 | 45.40 | 5.56 | Medicated (n = 31) | 0 | 1.5T/8 mm | *p* < 0.05 (FWE) | MDD < HC: R mPFC, R superior OFC |
| Opel et al. 2019 | 506 (308) | 358 (178) | 49.14 | 52.57 | 6.73 | Medicated (n = 441) | Comorbidity index: 1.65 ± 0.81 | 3T/8 mm | *p* < 0.05 (FWE) | MDD < HC: R Insula/STG/MTG, L SFG/MFG, R MFG/IFG |
| Pannekoek et al. 2014 | 26 (23) | 26 (23) | 15.40 | 14.70 | NA | Drug naïve | 18 anxiety disorder; 5 ADHD | 3T/7 mm | *p* < 0.05 (permutation) | -- |
| Qiu et al. 2016 | 12 (8) | 15 (10) | 34.40 | 33.70 | NA | Drug naïve | 0 | 3T/8 mm | *p* < 0.001 (uncorr) | MDD < HC: R Cingulate gyrus |

| | | | | | | | | | | |
|---|---|---|---|---|---|---|---|---|---|---|
| Redlich et al. 2014 | 58 (36) | 58 (37) | 37.60 | 37.70 | 10.98 | Medicated (n = 52) | 35 anxiety disorder | 3T/8 mm | $p < 0.05$ (AlphaSim) | MDD < HC: L/R Hippocampus, FFG, Lingual gyrus, L Supramarginal gyrus, IPL, L/R MCC, ACC, mPFC, SMA, R Middle frontal gyrus, SFG, L/R Precunues, L MTG/STG, L Caudate |
| Redlich et al. 2018 | 20 (15) | 21 (12) | 16.00 | 16.57 | 2.63 | Medicated (n = 8) | 0 | 3T/6 mm | $p < 0.05$ (FWE) | -- |
| Rodríguez-Cano et al. 2014 | 32 (20) | 64 (38) | 48.68 | 46.03 | 10.94 | Medicated (n = 28) | 2 AUD; 2 AG; 1 AD; 1 anxiety disorder; 1 dysthymia | 1.5T/4 mm | $p < 0.05$ (FWE) | MDD < HC: L FFG |

| | | | | | | | | | | |
|---|---|---|---|---|---|---|---|---|---|---|
| Salvadore et al. 2011 | 58 (37) | 107 (60) | 38.80 | 36.20 | 18.40 | Medicated (n = 44) | 0 | 3T/11 mm | *p* < 0.05 (FWE) | MDD < HC: L/R SFG, L MFG<br>MDD > HC: R Cerebellum, R Rolandic operculum, R SFG, R Precuneus, L IFG, R Amygdala; |
| Scheuerecker et al. 2010 | 13 (3) | 15 (5) | 37.90 | 35.50 | 4.36 | Drug naïve | 0 | 3T/8 mm | *p* < 0.001 (uncorr) | MDD < HC: L IFG, L Inferior frontal operculum, L MTG, R Postcentral gyrus, L IPL, L STG, L Postcentral gyrus, L Rolandic operculum, R IOG, L IFG, ITG |

| | | | | | | | | | | |
|---|---|---|---|---|---|---|---|---|---|---|
| Shad et al. 2012 | 22 (10) | 22 (11) | 15.00 | 16.00 | NA | Medicated (n = 4) | 2 anxiety disorder; 3 ADHD | 1.5T/8 mm | $p < 0.05$ (FWE) | MDD < HC: R IFG, L/R MFG, R SFG, L/R Caudate, R STG, L Thalamus, L/R Cerebellum |
| Sprengelmeyer et al. 2011 | 17 (9) | 21 (12) | 45.60 | 42.00 | NA | Medicated (n = 17) | 0 | 1.5T/8 mm | $p < 0.05$ (Monte Carlo) | MDD < HC: L Insula, R PHG, L MTG, R SFG, L FFG, R Amygdala |
| Stratmann et al 2014 | 132 (76) | 132 (74) | 37.86 | 37.82 | 7.78 | Medicated (n = 126) | 41 anxiety disorder | 3T/8 mm | $p < 0.05$ (AlphaSim) | MDD < HC: R Insula, L SPL, L/R STG, L PHG |
| Straub et al. 2019 | 60 (48) | 43 (38) | 17.30 | 17.62 | NA | Medicated (n = 20) | 18 unspecified | 3T/6 mm | $p < 0.05$ (FWE) | MDD > HC: R dlPFC |
| Treadway et al. 2009 | 19 (10) | 19 (10) | 35.20 | 30.30 | 12.90 | NA | 7 anxiety disorder | 3T/12 mm | $p < 0.05$ (FWE) | -- |
| Ueda et al. 2016 | 30 (13) | 48 (13) | 44.30 | 41.20 | NA | NA | 0 | 3T/8 mm | $p < 0.05$ (FWE) | MDD < HC: STG |

| | | | | | | | | | | |
|---|---|---|---|---|---|---|---|---|---|---|
| Wagner et al. 2011 | 30 (25) | 30 (25) | 37.55 | 35.10 | 5.59 | NA | 0 | 1.5T/12 mm | $p < 0.05$ (FWE) | MDD < HC: L/R Nucleus caudatus, R IFG, R Subgenual cortex, L/R Hippocampus/amygdala, L OFC, L SFG |
| Wehry et al. 2015 | 14 (11) | 41 (27) | 14.00 | 13.00 | NA | NA | 5 ADHD; 1 disruptive behavior | 4T/8 mm | $p < 0.001$ (uncorr) | MDD > HC: R MFG, L Precuneus, R Thalamus, R Caudate |
| Yang et al. 2017 | 35 (35) | 23 (23) | 44.54 | 39.09 | 2.68 | Drug naïve | NA | 3T/8 mm | $p < 0.005$ (AlphaSim) | MDD < HC: L/R MFG, L/R Insula, L/R Putamen, R Amygdala, R PHG, L Lingual gyrus, Cerebellum |
| Yüksel et al. 2018 | 37 (20) | 54 (21) | 37.90 | 35.90 | NA | Medicated (n = 28) | 13 OCD, somatoform | 3T/NA | $p < 0.05$ (FWE) | -- |

| | | | | | | | | | | |
|---|---|---|---|---|---|---|---|---|---|---|
| | | | | | | | or personality disorder | | | |
| Zhao et al. 2017 | 37 (12) | 41 (15) | 26.70 | 27.10 | 2.00 | Drug naïve | 0 | 3T/10 mm | $p < 0.001$ (FDR) | MDD > HC: R Temporal pole; MDD < HC: L/R OFC, L/R Putamen, L/R Thalamus, L/R MFG, L Cuneus |
| Zhou et al. 2018 | 144 (84) | 111 (58) | 28.17 | 27.63 | NA | Drug naïve | 0 | 3T/8 mm | $p < 0.05$ (AlphaSim) | MDD < HC: R SOG |

Abbreviations: ACC, anterior cingulate cortex; AD, adjustment disorder with mixed disturbance of emotions and conduct; ADHD, attention deficit hyperactivity disorder; AG, agoraphobia; AUD, alcohol use disorder; corr, corrected; dlPFC, dorsolateral prefrontal cortex; dmPFC, dorsomedial prefrontal cortex; FDR, false discovery rate; FFG, fusiform gyrus; FWE, family-wise error; FWHM, full width at half maximum; GRF, Gaussian random field; HC, healthy controls; IFG, inferior frontal gyrus; IOG, inferior occipital gyrus; IPL, inferior parietal lobule; ITG, inferior temporal gyrus; L, left hemisphere; MCC, medial cingulate cortex; MDD, major depressive disorder; MFG, medial frontal gyrus; MOG, middle occipital gyrus; mPFC, medial prefrontal cortex; MTG, middle temporal gyrus; NA, not available; OCD, obsessive-compulsive disorder; OFC, orbitofrontal cortex; PCC, posterior cingulate cortex; PHG, parahippocampal gyrus; R, right hemisphere; SFG, superior frontal gyrus; sgACC, subgenual anterior cingulate cortex; SMA, supplementary motor area; SOG, superior occipital gyrus; SPL, superior parietal lobule; STG, superior temporal gyrus; uncorr, uncorrected; VBM, voxel-based morphometry; vmPFC ventromedial prefrontal cortex.

* Studies that provided original whole-brain *t*-maps.

**Table S4.** Whole-brain meta-analysis results for VBM studies in GAD, FAD and MDD at $p < 0.0025$, uncorrected.

| MNI coordinates | SDM Z-Score | P *value* | Voxels | Regions | BA | Egger's bias | Egger's p value |
|---|---|---|---|---|---|---|---|
| **GAD > HC** | | | | | | | |
| 4, -28, 62 | 3.411 | 0.00032 | 18 | R paracentral lobule | 4 | -0.37 | 0.832 |
| **GAD < HC** | | | | | | | |
| -44, -8, 8 | -3.951 | 0.000039 | 411 | L insula, rolandic operculum, heschl gyrus, STG, supramarginal gyrus | 48 | 0.45 | 0.806 |
| -32, 24, -10 | -3.973 | 0.000035 | 247 | L IFG, temporal pole, insula | 38/47 | -0.41 | 0.809 |
| -8, -24, 12 | -3.694 | 0.00011 | 22 | L thalamus | -- | 0.26 | 0.878 |
| 6, -58, 10 | -3.117 | 0.00091 | 12 | R lingual gyrus | 17 | 0.77 | 0.697 |
| 44, -38, 50 | -3.177 | 0.00074 | 10 | R IPG | 2 | 0.28 | 0.873 |
| **FAD > HC** | | | | | | | |
| 12, -70, -12 | 3.626 | 0.000144 | 26 | R lingual gyrus | 18 | -0.01 | 0.993 |
| -14, -80, -12 | 3.331 | 0.000433 | 20 | L lingual gyrus | 18 | 0.05 | 0.942 |
| **SAD > HC** | | | | | | | |
| -56, -20, 22 | 3.514 | 0.000220 | 147 | L postcentral gyrus, STG | 48 | 0.26 | 0.797 |

| | | | | | | | |
|---|---|---|---|---|---|---|---|
| 48, -46, 34 | 3.476 | 0.000255 | 91 | R superior longitudinal fasciculus, angular gyrus | 39/40 /48 | 0.60 | 0.602 |
| 26, -50, 58 | 3.484 | 0.000246 | 66 | R SPG | 5/7 | 0.11 | 0.909 |
| 24, 56, 12 | 3.641 | 0.000136 | 57 | R SFG, dorsolateral | 10 | 0.05 | 0.959 |
| -20, -32, 62 | 3.885 | 0.000051 | 35 | L postcentral/precentral gyrus | 3 | -0.08 | 0.933 |
| 56, -26, 48 | 3.342 | 0.000416 | 20 | R postcentral gyrus, supramarginal gyrus | 2/3 | 0.03 | 0.975 |
| -28, -42, -46 | 3.574 | 0.000176 | 19 | L cerebellum, | -- | -0.07 | 0.944 |
| -8, 52, 12 | 3.380 | 0.000363 | 16 | L SFG, medial | 10 | -0.31 | 0.74 |
| 14, -68, -10 | 3.145 | 0.000829 | 11 | R lingual gyrus | 18 | 0.15 | 0.874 |
| -12, -80, -12 | 3.265 | 0.000547 | 11 | L lingual gyrus | 18 | 0.56 | 0.587 |
| **PD < HC** | | | | | | | |
| 38, 0, 8 | -3.784 | 0.000077 | 63 | R Insula | 48 | -0.79 | 0.488 |
| 48, -2, -10 | -3.200 | 0.000687 | 12 | R STG | 48 | -0.70 | 0.494 |
| -36, 10, -22 | -3.037 | 0.001195 | 12 | L temporal pole, STG | 38 | 0.09 | 0.928 |
| **MDD < HC** | | | | | | | |
| 46, -2, 4 | -4.339 | 0.0000071 53 | 877 | R insula, MTG, STG, rolandic operculum | 48/21 /22 | -0.37 | 0.405 |
| 2, 36, -10 | -5.012 | 0.0000002 98 | 722 | L/R SFG, medial orbital, L/R ACG | 11/10 /32 | -0.54 | 0.300 |

| | | | | | | | |
|---|---|---|---|---|---|---|---|
| 10, 6, 40 | -3.709 | 0.0001041 29 | 118 | L/R MCG | 32/24 | -0.31 | 0.591 |
| 36, -30, -14 | -3.115 | 0.0009192 23 | 10 | R PHG | 20 | -0.59 | 0.371 |

Abbreviations: ACG, anterior cingulate/paracingulate gyri; BA, Brodmann area; FAD, fear-related anxiety disorder; GAD, generalized anxiety disorder; HC, healthy controls; IFG, inferior frontal gyrus; IPG, inferior parietal gyri; ITG, inferior temporal gyrus; L, left hemisphere; MCG, median cingulate/paracingulate gyri; MDD, major depressive disorder; MNI, Montreal Neurological Institute; MTG, middle temporal gyrus; PHG, parahippocampal gyrus; R right hemisphere; SDM, seed-based d mapping; SFG, superior frontal gyrus; SPG, superior parietal gyri; STG, superior temporal gyrus; VBM, voxel-based morphometry.

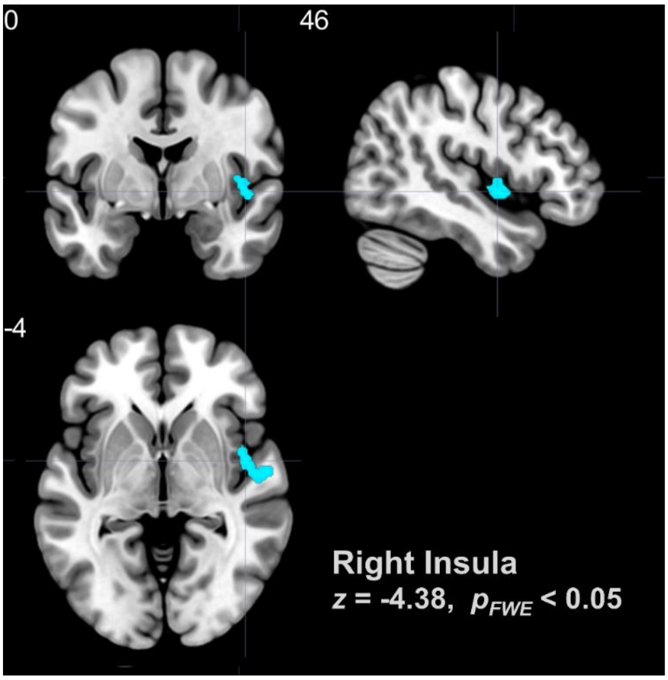

**Figure. S1**. Brain regions showing decreased GMV of patient vs healthy controls for studies pooled across GAD, FAD and MDD. Clusters were displayed at TFCE based FWE corrected $p < 0.05$.
